# Cross-cohort generalizability of machine learning models for early-stage node-negative breast cancer relapse prediction

**DOI:** 10.64898/2026.10.02.26364592

**Authors:** Cagri Ozdemir, Sangeeta Gupta

## Abstract

**Background:** Adjuvant therapy decision in early-stage node-negative breast cancer remains one of the most challenging problems in oncology. Although most patients in this population achieve favorable outcomes with endocrine therapy alone, a subset remains at risk of relapse and may benefit from chemotherapy. Accurate identification of high-risk and low-risk patients is therefore essential to prevent both undertreatment and overtreatment.

**Methods:** In this study, machine learning models were assessed using MSK-IMPACT (n=645) and METABRIC (n=503) cohorts comprising node-negative stage I-II breast cancer patients. Three experimental settings were evaluated: training on MSK-IMPACT and testing on METABRIC, the reverse direction, and five-fold cross-validation on the combined cohort. Sensitivity analysis and data harmonization were applied to identify and mitigate cohort-specific bias. Survival analysis, SHapley Additive exPlanations (SHAP) interpretability, and ablation analyses were performed to validate clinical relevance and characterize feature contributions.

**Results:** The combined cohort model achieved an area under the receiver operating characteristic curve (AUC) of 0.788*±*0.028 across five-fold cross-validation, while bidirectional cross-cohort validation yielded AUC of 0.741 when testing on MSK-IMPACT and AUC of 0.714 when testing on METABRIC. Survival validation on METABRIC out-of-fold predictions from the combined cohort model showed hazard ratios of 3.01 (95% CI: 2.28–3.98) for disease-free survival and 2.35 (95% CI: 1.79–3.09) for overall survival, consistent across Luminal A, Luminal B, and Triple-Negative subtypes.

**Conclusion:** Validated across two geographically independent cohorts, the proposed model enables cost-effective, gene expression–free identification of patients at high-risk of distant relapse and may facilitate adjuvant chemotherapy decision-making in early-stage node-negative breast cancer.

## 1. Introduction

Node-negative early-stage breast cancer presents one of the most challenging risk stratification scenarios in clinical oncology. Despite the absence of lymph node involvement, which is generally considered a favorable prognostic indicator, a significant proportion of node-negative patients remain at risk of distant relapse and may therefore benefit from adjuvant chemotherapy [1, 2]. However, current tools are insufficient to reliably identify this subgroup, leading to both overtreatment and undertreatment [3].

Several prognostic tools have been developed to support clinical decision making in breast cancer. Gene expression-based assays such as Oncotype DX [4], Mamma Print [5], and PAM50 [6] have demonstrated prognostic utility. However, these assays require dedicated molecular profiling platforms and remain costly, limiting accessibility in many low-resource health-care settings [7, 8]. Clinically based tools such as PREDICT, which rely on routinely collected clinicopathological variables, offer broad accessibility but exhibit limited discriminatory performance. In node-negative patients, PREDICT has reported an area under the receiver operating characteristic curve (AUC) of only 0.65 (95% CI: 0.62–0.68), with particularly poor performance in estrogen receptor (ER)-negative tumors (AUC of 0.56) [9]. This performance gap highlights the urgent need for approaches that integrate genomic data while remaining deployable in standard clinical sequencing workflows.

Machine learning models have also shown promise for cancer prognosis prediction; however, most published studies validate their approaches within a single institutional cohort using train–test splits derived from the same data source [10, 11, 12, 13]. This practice can lead to shortcut learning, in which models rely on cohort-specific characteristics that are unique to a particular dataset instead of learning biologically meaningful patterns that generalize to new patient populations [14]. The consequences of such bias become apparent only when models are tested in truly independent external cohorts, a validation step that remains uncommon in the breast cancer machine learning literature. Furthermore, in predictive modeling, a crucial trade-off arises: Do we merely desire the prediction, or are we interested in understanding the rationale behind it? [15]. Because each clinicopathological and genomics feature reflects a distinct aspect of the underlying biology, quantifying their relative importance may enhance understanding of breast cancer recurrence and help advance precision medicine in oncology.

To address these limitations, we developed a multi-cohort, interpretable machine learning framework for relapse prediction in node-negative stage I–II breast cancer. The framework integrates clinicopathological variables with genomic data routinely generated by standard clinical sequencing workflows and evaluates its generalizability across two geographically and institutionally independent cohorts: the Memorial Sloan Kettering Cancer Center (MSK-IMPACT) [16] from the United States and the Molecular Taxonomy of Breast Cancer International Consortium (METABRIC) [17] from the United Kingdom and Canada. We further assessed the clinical utility of the machine learning model by evaluating its ability to stratify patients into groups with significantly different survival outcomes and by examining performance across molecular subtypes. Model predictions were interpreted using SHapley Additive exPlanations (SHAP) analysis [18] and feature ablation studies to identify the most influential predictors. Data harmonization and heterogeneous multi-cohort training substantially improved model robustness and generalizability. The resulting framework offers a gene expression-free approach to relapse risk stratification in node-negative early-stage breast cancer. To promote reproducibility and facilitate future research, the final model trained on the combined cohort has been made publicly available.

## 2. Methods

### 2.1. Data preprocessing and cohort harmonization

This multi-cohort study integrated genomic and clinicopathological data from two publicly available breast cancer datasets: MSK-IMPACT [16] and METABRIC [17]. Both datasets were accessed through cBioPortal (https://www.cbioportal.org/). Patients with stage I-II, no regional lymph node involvement, and no distant metastasis at the diagnosis were included. The relapse group comprised patients with documented distant metastatic recurrence following an initial disease-free period. Recurrence labels were defined using disease recurrence status fields available in each cohort (total n=548; MSK-IMPACT n=316, METABRIC n=232). The non-relapse group comprised patients who remained disease-free through their last follow-up. Non-relapse labels were confirmed using cohort-specific criteria (total n=600; MSK-IMPACT n=329, METABRIC n=271). The inclusion criteria were applied as follows: (i) absence of local and metastatic recurrence; (ii) minimum disease-free survival of 60 months; (iii) confirmed living status at last follow-up. For MSK-IMPACT, the minimum disease-free survival criterion of 60 months was not applied because prospective clinical sequencing cohorts typically have shorter follow-up durations. After applying the inclusion criteria, 1,148 patients were eligible for analysis (MSK-IMPACT n=645, METABRIC n=503).

For each patient, a total of 14 variables were collected comprising six clinicopathological features and eight genomic features. Age, menopausal status, histological grade, estrogen receptor (ER), progesterone receptor (PR), and human epidermal growth factor receptor 2 (HER2) status were used as clinicopathological features. Features were selected based on established clinical relevance and prior biological evidence for breast cancer prognosis. Variable descriptions and encoding schemes are provided in Supplementary Table S1. Genomic features included total single nucleotide variant (SNV) counts, Combined Annotation Dependent Depletion (CADD) pathogenicity scores [19], and raw copy number alterations (CNAs). CADD scores were retrieved using the CADD web server https://cadd.gs.washington.edu/, and the maximum score per gene per sample was retained as a representative pathogenicity feature. SNV and CADD features were derived from TP53 and PIK3CA, two well-established breast cancer driver genes [20]. CNA features included CCND1 and MDM4, which were selected based on their statistical association with disease recurrence across the cohorts and their functional relevance to TP53 signaling. Tumor mutation burden (TMB) and fraction genome altered (FGA) were also included as genomic data. TMB values were harmonized across cohorts using a rank-based inverse normal transformation. Within each cohort, TMB values were ranked and transformed to a standard normal distribution using the quantile function, thereby reducing platform-specific scale differences. This approach has been previously validated for cross-panel TMB harmonization [21]. FGA values were inherently normalized to a 0–1 scale in both cohorts and were utilized directly as continuous features without further processing. In MSK-IMPACT cohort, missing data rates ranged from 0% to 16.1%, with the highest missingness observed for histological grade. In the METABRIC cohort, missing data rates ranged from 0% to 13.7%, with the highest missingness observed for menopausal status, PR status, and HER2 status (Supplementary Table S2). Missing values were handled using k-nearest neighbor (kNN) imputation (k = 5), where missing values were estimated using the weighted mean of the five most similar patients. Binary features (ER status, PR status, HER2 status, and menopausal status) were rounded to the nearest integer (0 or 1), and histological grade was rounded to the nearest integer and clipped to the valid range of 1–3 to ensure clinical validity of imputed values. Imputation was performed independently within each cohort to prevent data leakage between the discovery and validation sets.

### 2.2. Predictive modeling and evaluation

The predictive performance of four machine learning models were evaluated for the task of breast cancer relapse prediction: Random Forest (RF), Gradient Boosting (GB), Adaptive Boosting (AdaBoost), and eXtreme Gradient Boosting (XGBoost). Each model’s performance was assessed using AUC, area under the precision-recall curve (AUCPR), F1-score, recall, and precision under three experimental settings: (i) training on MSK-IMPACT and testing on METABRIC (cross-cohort validation); (ii) training on METABRIC and testing MSK-IMPACT (reverse cross-cohort generalization); and (iii) training and evaluating on the combined cohort using 5-fold cross validation repeated five times (25 evaluations in total). To evaluate the impact of cohort-specific features on model performance, a sensitivity analysis was conducted by excluding features with significant distributional differences between cohorts and repeating all three experiments. For the cross-cohort (experiment i) and reverse cross-cohort (experiment ii), hyperparameters were optimized independently within each experimental setting using inner cross-validation on the training data only, ensuring no information from the test cohort influenced model selection. For the combined cohort analysis (experiment iii), hyperparameter tuning was performed using repeated 5-fold cross-validation (25 outer runs). Within each outer training fold, hyperparameters were optimized using inner cross-validation, and the best-performing configuration from each run was recorded.

A final predictive model was developed for clinical deployment using all samples from the combined cohort (MSK-IMPACT and METABRIC). The final hyperparameter set was selected as the mode (most frequently selected configuration) across the 25 outer cross-validation runs, representing the most stable solution across resampling variability. The model was subsequently retrained using the entire combined cohort with the selected hyperparameters. The resulting model can directly generate relapse risk predictions for new patients using their clinical and genomic features and is publicly available for clinical decision support. Optimal hyperparameters for each experimental setting were reported in Supplementary Table S3.

### 2.3. Survival analysis and prognostic evaluation

Survival analysis was performed to evaluate the prognostic significance of model-derived relapse risk scores under two settings. First, using the best-performing model from the combined cohort analysis (experiment iii), out-of-fold relapse probability scores were generated for all patients through stratified 5-fold cross-validation, ensuring that each prediction was obtained from a model that had not been trained on the corresponding patient sample. Second, to provide a fully independent cross-cohort survival validation, the best-performing model was trained on the full MSK-IMPACT cohort and applied to all METABRIC patients. In both settings, patients were stratified into high-risk and low-risk groups using a probability threshold of 0.5. Kaplan-Meier survival curves were constructed for disease-free survival (DFS) and overall survival (OS), and differences between groups were assessed using the log-rank test. Associations between predicted risk groups and survival outcomes were further evaluated using univariate Cox proportional hazards regression, with hazard ratios (HRs) and 95% confidence intervals reported. Survival analyses were restricted to the METABRIC cohort, as the MSK-IMPACT cohort exhibited a high proportion of censored non-relapse patients with insufficient follow-up time for meaningful survival analysis. DFS and OS analyses were performed on non-overlapping patient subsets after excluding samples with missing survival data, retaining 502 patients for DFS and 434 patients for OS.

### 2.4. Feature importance and ablation analysis

To assess the contribution of individual features to model predictions, SHAP values were computed for the best-performing model, which was trained on the full combined cohort [18]. To further evaluate the relative contribution of different feature categories, an ablation analysis was performed by training and evaluating the combined cohort model using three feature subsets separately: genomic features only, clinicopathological features only, and the full combined feature set. All three experimental settings were repeated for each feature subset, and the resulting AUC values were compared to quantify the contribution of each feature category to predictive performance.

## 3. Results

### 3.1. Cohort characteristics

A total of 1,148 patients meeting the inclusion criteria were identified across two cohorts. MSK-IMPACT [16] contributed 645 patients, including 316 relapse and 329 non-relapse cases, METABRIC [17] contributed 503 patients, including 232 relapse and 271 non-relapse cases. After preprocessing, imputation, and harmonization, clinicopathological and genomic characteristics of the study cohorts were summarized in Table 1. Continuous variables were presented as mean *±* standard deviation (sd) and were compared between relapse and non-relapse groups using the Mann–Whitney U test [22]. Binary variables were presented as counts and percentages and were compared using the Chi-square test [23]. Relapse and non-relapse groups were reasonably balanced in both MSK-IMPACT and METABRIC, providing a suitable foundation for predictive modeling. Several variables differed significantly between relapse and non-relapse groups, including histological grade, TMB, FGA, TP53 SNV counts, TP53 CADD pathogenicity scores, CCND1 CNAs, and MDM4 CNAs. Both cohorts included representation of all major breast cancer molecular subtypes, defined using immunohistochemistry (IHC)-based surrogate criteria based on ER, PR, and HER2 status [24]. In MSK-IMPACT cohort, Luminal A (ER+ / PR+ / HER2*−*) was the predominant subtype in both relapse (62.7%) and non-relapse (70.5%) groups, followed by Luminal B (ER+ / PR*−* / HER2*−* or ER+ / any / HER2+; 20.9% and 13.1%), Triple-Negative (ER*−* / PR*−* / HER2*−*; 12.3% and 12.8%), and other subtypes (4.1% and 3.6%). A similar distribution was observed in METABRIC, with Luminal A accounting for 53.4% and 59.0% of relapse and non-relapse patients respectively, followed by Luminal B (25.4% and 20.7%), Triple-Negative (15.1% and 14.8%), and other subtypes (6.0% and 5.5%).

**Table 1:** Clinicopathological and genomic characteristics of relapse and non-relapse patients in MSK-IMPACT and METABRIC cohorts. Statistical significance is indicated as follows: * p < 0.05, ** p < 0.01, *** p < 0.001, **** p < 0.0001.

| Feature | MSK-IMPACT |  |  | METABRIC |  |  |
| --- | --- | --- | --- | --- | --- | --- |
|  | Relapse | Non-relapse | p-value | Relapse | Non-relapse | p-value |
| Age, mean (sd) | 50 ± 10 | 59 ± 12 | **** | 59 ± 12 | 59 ± 12 | 0.553 |
| Menopausal status, n (%) |  |  | **** |  |  | 0.254 |
| Pre | 184 (58.2%) | 115 (35.0%) |  | 63 (27.2%) | 61 (22.5%) |  |
| Post | 132 (41.8%) | 214 (65.0%) |  | 169 (72.8%) | 210 (77.5%) |  |
| Histological grade, n (%) |  |  | **** |  |  | *** |
| grade I | 0 (0.0%) | 0 (0.0%) |  | 14 (6.0%) | 43 (15.9%) |  |
| grade II | 75 (23.7%) | 153 (46.5%) |  | 93 (40.1%) | 143 (52.8%) |  |
| grade III | 241 (76.3%) | 176 (53.5%) |  | 125 (53.9%) | 85 (31.4%) |  |
| ER status, n (%) |  |  | 1.0 |  |  | 0.826 |
| positive | 264 (83.5%) | 275 (83.6%) |  | 183 (78.9%) | 216 (79.7%) |  |
| negative | 52 (16.5%) | 54 (16.4%) |  | 49 (21.1%) | 55 (20.3%) |  |
| PR status, n (%) |  |  | 0.092 |  |  | 0.465 |
| positive | 221 (69.9%) | 250 (76.0%) |  | 136 (58.6%) | 168 (62.0%) |  |
| negative | 95 (30.1%) | 79 (24.0%) |  | 96 (41.4%) | 103 (38.0%) |  |
| HER2 status, n (%) |  |  | * |  |  | * |
| positive | 32 (10.1%) | 18 (5.5 %) |  | 26 (11.2%) | 16 (5.9%) |  |
| negative | 284 (89.9%) | 311 (94.5%) |  | 206 (88.8%) | 255 (94.1%) |  |
| FGA, mean (sd) | 0.38 ± 0.22 | 0.19 ± 0.18 | **** | 0.32 ± 0.18 | 0.20 ± 0.15 | **** |
| TMB, mean (sd) | 0.26 ± 0.98 | -0.22 ± 0.89 | **** | 0.11 ± 0.97 | -0.09 ± 0.99 | ** |
| TP53 SNVs mutated, n (%) | 105 (33.2%) | 74 (22.5%) | *** | 65 (28.0%) | 47 (17.3%) | *** |
| TP53 CADD, mean (sd) | 9.41 ± 13.68 | 6.81 ± 12.92 | * | 8.14 ± 13.39 | 4.83 ± 10.81 | ** |
| PIK3CA SNVs mutated, n (%) | 113 (35.8%) | 152 (46.2%) | *** | 96 (41.4%) | 122 (45.0%) | *** |
| PIK3CA CADD, mean (sd) | 9.16 ± 12.47 | 12.06 ± 13.24 | ** | 10.93 ± 13.22 | 11.45 ± 12.89 | 0.885 |
| CCND1 CNAs, mean (sd) | 0.45 ± 0.84 | 0.22 ± 0.63 | *** | 0.56 ± 0.92 | 0.30 ± 0.69 | *** |
| MDM4 CNAs, mean (sd) | 0.11 ± 0.46 | 0.02 ± 0.19 | *** | 0.92 ± 0.78 | 0.68 ± 0.80 | *** |
| Luminal A, n (%) | 198 (62.7%) | 232 (70.5%) | - | 124 (53.4%) | 160 (59.0%) | - |
| Luminal B, n (%) | 66 (20.9%) | 43 (13.1%) | - | 59 (25.4%) | 56 (20.7%) | - |
| Triple-Negative, n (%) | 39 (12.3%) | 42 (12.8%) | - | 35 (15.1%) | 40 (14.8%) | - |
| Other subtypes, n (%) | 13 (4.1%) | 12 (3.6%) | - | 14 (6.0%) | 15 (5.5%) | - |

### 3.2. Predictive performance and cross-cohort generalizability

Age and menopausal status distributions differed significantly between relapse and non-relapse patients in MSK-IMPACT, with p< 0.0001 for both features, however they showed no significant differences in METABRIC, where age yielded p = 0.553 and menopausal status p = 0.254 (Table 1). To assess whether these features introduced bias in cross-cohort generalization, a sensitivity analysis was performed by repeating experiments after excluding age and menopausal status from the feature set. Each machine learning model performance was assessed using two cross-cohort experiments: training on MSK-IMPACT and testing on METABRIC, and training on METABRIC and testing on MSK-IMPACT. In addition, model performance was evaluated using repeated 5-fold cross-validation on the combined cohort (25 evaluations in total). Table 2 summarizes AUC and AUCPR performance of all models across the three experimental settings, where cross-cohort experiments were reported after excluding the cohort-specific features to minimize cohort-specific bias, while the combined cohort model retained all features to leverage the full heterogeneity of the pooled dataset. The sensitivity analysis revealed that excluding age and menopausal status had a differential impact across experimental settings. In experiment (i) (train MSK *→* test METABRIC), performance improved after exclusion across all models. For RF, GBM, AdaBoost, and XGBoost, AUC increased from 0.677 to 0.714, 0.637 to 0.669, 0.645 to 0.674, and 0.630 to 0.668, respectively, indicating improved cross-cohort generalization following removal of cohort-specific features (Fig. 1a). In contrast, performance in experiment (ii) (train METABRIC → test MSK) decreased from 0.767 to 0.741, 0.741 to 0.700, 0.733 to 0.717, and 0.728 to 0.688, respectively (Fig. 1b). For the combined cohort model (experiment (iii)), including age and menopausal status led to a statistically significant performance increase in RF, GBM, AdaBoost, and XGBoost (Fig. 1c). The AUC values increased from 0.767, 0.766, 0.759, and 0.767 to 0.781, 0.785, 0.784, and 0.788 respectively. In cross-cohort experiments, RF achieved the highest AUC and AUCPR. Although XGBoost achieved the highest AUC in the combined cohort setting, RF demonstrated the most consistent cross-cohort generalization across both validation directions and produced more stable and interpretable SHAP values. RF was therefore selected as the final model. Full metrics including recall, precision, F1, and specificity are provided in Supplementary Table S4.

**Figure 1:**
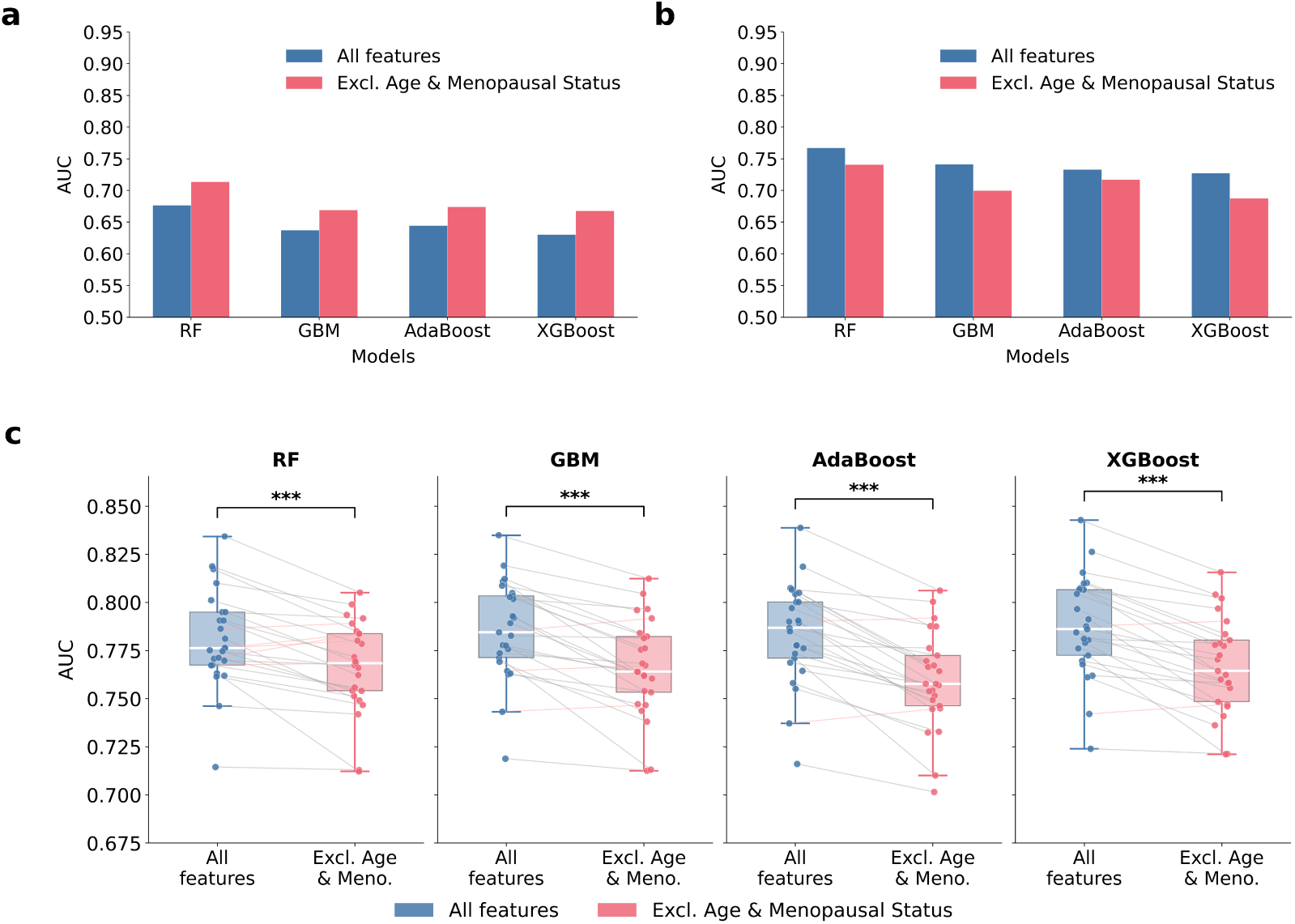
Predictive performance of machine learning models across experimental settings and sensitivity analysis. **a** AUC comparison for experiment (i) (Train MSK-IMPACT → Test METABRIC) showing model performance with all features versus after excluding cohort-specific features (age and menopausal status). **b** AUC comparison for experiment (ii) (Train METABRIC → Test MSK-IMPACT) under the same two feature conditions. **c** Paired comparison of AUC distributions from 25 repeated cross-validation runs (experiment iii) with and without age and menopausal status, showing the impact of cohort-specific feature exclusion on combined cohort performance. Statistical significance between conditions in was assessed using the Wilcoxon signed-rank test; *** p<0.001.

**Table 2:** Predictive performance of machine learning models for breast cancer relapse prediction across experimental settings. Metrics include area under the receiver operating characteristic curve (AUC) and area under the precision–recall curve (AUCPR). For the combined cohort setting, results are reported as mean ± standard deviation across 5-fold cross-validation repeated five times (25 runs). The best performance per metric and experimental setting is highlighted in bold. MSK: MSK-IMPACT cohort; META: METABRIC cohort.

| Experiments | RF |  | GBM |  | AdaBoost |  | XGBoost |  |
| --- | --- | --- | --- | --- | --- | --- | --- | --- |
|  | AUC | AUCPR | AUC | AUCPR | AUC | AUCPR | AUC | AUCPR |
| Train MSK → Test META | **0.714** | **0.663** | 0.669 | 0.603 | 0.674 | 0.627 | 0.668 | 0.631 |
| Train META → Test MSK | **0.741** | **0.689** | 0.700 | 0.646 | 0.717 | 0.675 | 0.688 | 0.631 |
| Combined cohort | 0.781±0.024 | 0.758±0.029 | 0.785±0.025 | 0.775±0.029 | 0.784±0.026 | 0.772±0.031 | **0.788±0.026** | **0.778±0.028** |

### 3.3. Survival analysis and prognostic validation

To validate the prognostic significance of the model-derived risk scores, Kaplan-Meier survival analysis and univariate Cox proportional hazards regression were performed on METABRIC patients. These analyses used out-of-fold test sets predictions from the combined cohort cross-validation with the RF classifier. Patients classified as high-risk by the model showed significantly worse DFS compared to those classified as low-risk ( log-rank p<0.0001, Fig. 2a). The median DFS was 85.6 months in the high-risk group, whereas the low-risk group did not reach the median. Cox regression further demonstrated that predicted high-risk status was significantly associated with shorter DFS, with a HR of 3.01 (95% CI: 2.28–3.98, p<0.0001). Similarly, patients classified as high-risk demonstrated significantly inferior OS, with a median OS of 129.8 months compared to 237.4 months in the low-risk group (log-rank p<0.0001, Fig. 2b). Cox regression confirmed a significant association between predicted high-risk status and worse OS, with a HR of 2.35 (95% CI: 1.79–3.09, p<0.0001).

**Figure 2:**
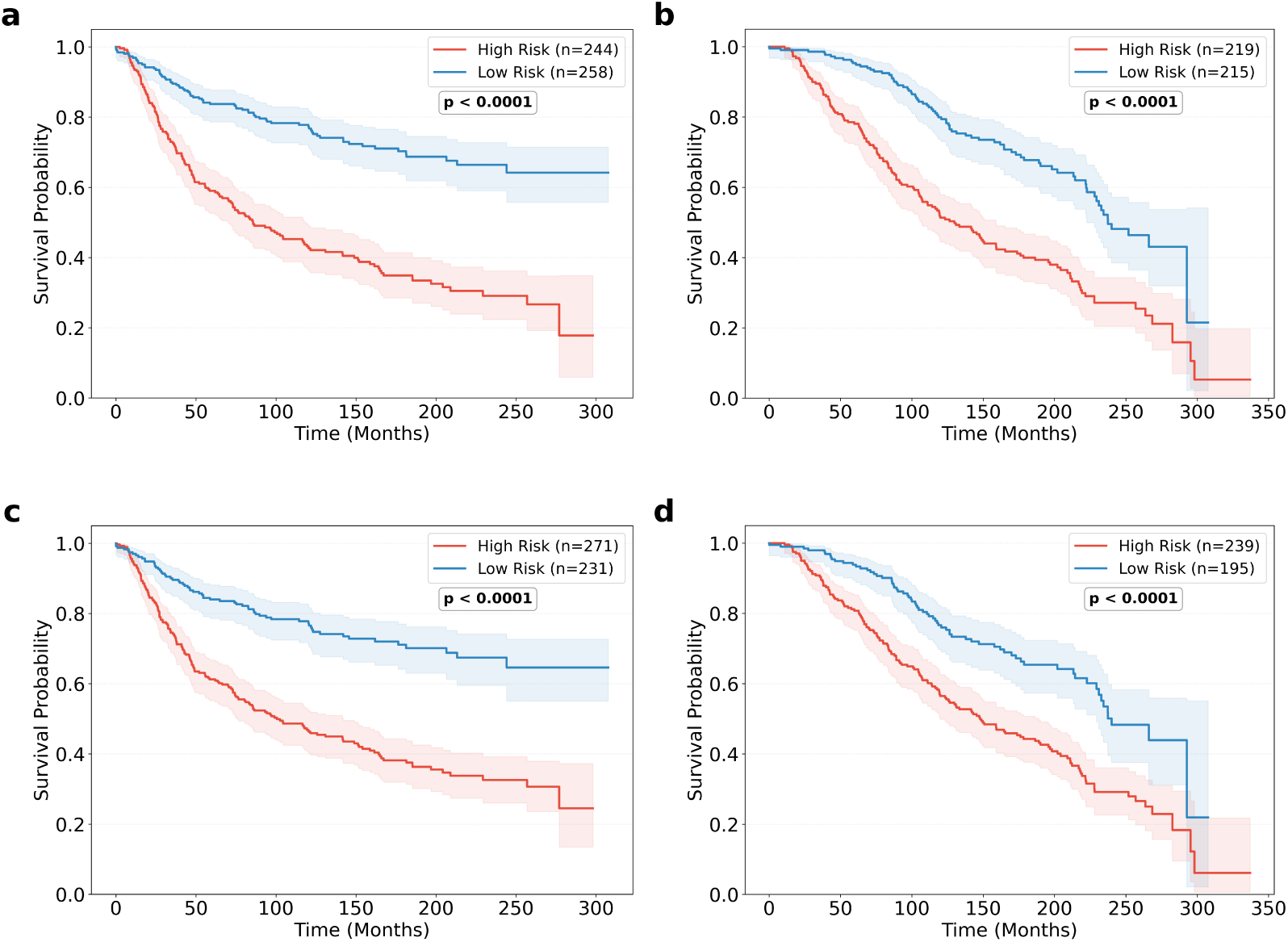
Kaplan-Meier survival curves for the model-derived risk stratification. **a** and **b** show disease-free survival (DFS) and overall survival (OS) based on out-of-fold test set predictions from the combined cohort cross-validation (RF trained on MSK-IMPACT + METABRIC, 5-fold CV). **c** and **d** show DFS and OS based on cross-cohort predictions (RF trained on MSK-IMPACT, tested on METABRIC), with age and menopausal status excluded to minimize cohort-specific bias. Patients were stratified into high-risk and low-risk groups using a probability threshold of 0.5. Shaded regions represent 95% confidence intervals.

To assess whether the prognostic value of the model generalized across cohorts, survival analyses were repeated using relapse risk scores generated by the RF model trained on the MSK-IMPACT cohort and applied to the independent METABRIC cohort (experiment i). Age and menopausal status were excluded to minimize cohort-specific bias. Despite being trained on an external cohort, the model remained significantly associated with both DFS and OS in METABRIC (both log-rank p<0.0001; Fig. 2c and 2d), with hazard ratios of 2.81 (95% CI: 2.10–3.75, p<0.0001) for DFS and 2.05 (95% CI: 1.55–2.72, p<0.0001) for OS. Similar to the combined cohort analysis, patients classified as high-risk experienced significantly poorer survival outcomes than those classified as low-risk. Notably, despite a moderate cross-cohort AUC of 0.714 and AUCPR of 0.663, the model retained strong prognostic discrimination in the independent METABRIC cohort.

Subtype-stratified survival analysis demonstrated consistent prognostic discrimination across all major molecular subtypes (Supplementary Fig. S1). For DFS, significant separation between high and low-risk groups was observed in Luminal A (HR=2.80, 95% CI: 1.96–3.99, p<0.0001), Luminal B (HR=3.15, 95% CI: 1.80–5.48, p<0.0001), and Triple-Negative subtypes (HR=4.41, 95% CI: 1.99–9.76, p<0.0001), with the strongest prognostic effect observed in Triple-Negative patients. The median DFS was not reached in the low-risk group across all subtypes, while high-risk patients showed median DFS of 99.1, 56.0, and 46.4 months for Luminal A, Luminal B, and Triple-Negative subtypes respectively. For OS, significant stratification was maintained across all subtypes, with hazard ratios of 1.51 (95% CI: 1.04–2.19, p=0.032) for Luminal A, 1.91 (95% CI: 1.15–3.18, p=0.013) for Luminal B, and 2.83 (95% CI: 1.42–5.67, p=0.003) for Triple-Negative patients. Notably, Triple-Negative patients showed the largest OS difference, with median OS of 65.5 months in the high-risk group compared to 292.7 months in the low-risk group.

### 3.4. Genomic and clinicopathological feature contribution analysis

SHAP analysis of the final combined model revealed that genomic features collectively accounted for 63.2% of the total mean absolute SHAP contribution, while clinicopathological features contributed 36.8% (Fig. 3a). Among individual features, FGA was the most important predictor, followed by histological grade, age, and TMB (Fig. 3b). The beeswarm plot illustrated both the magnitude and direction of feature contributions to model predictions (Fig. 3c). Features with positive SHAP values shifted predictions toward relapse, whereas features with negative SHAP values shifted predictions toward non-relapse. The color gradient further reflects feature values, enabling visualization of how high and low feature levels influenced prediction outcomes. Ablation analysis demonstrated that the full combined feature set significantly outperformed both clinicopathological features alone (AUC 0.674 *±* 0.036 vs. 0.781 *±* 0.024, p<0.001) and genomic features alone (AUC 0.756 *±* 0.024 vs. 0.781 *±* 0.024, p<0.001) (Fig. 3d). These findings indicate that genomic and clinicopathological variables provide complementary, non-redundant information, and that integrating both feature types yields superior predictive performance. To support individual-level clinical interpretability, the final model generates patient-specific relapse risk probabilities alongside SHAP-based waterfall plots, enabling clinicians to visualize the contribution of each feature to an individual patient’s prediction (Supplementary Fig. S2).

**Figure 3:**
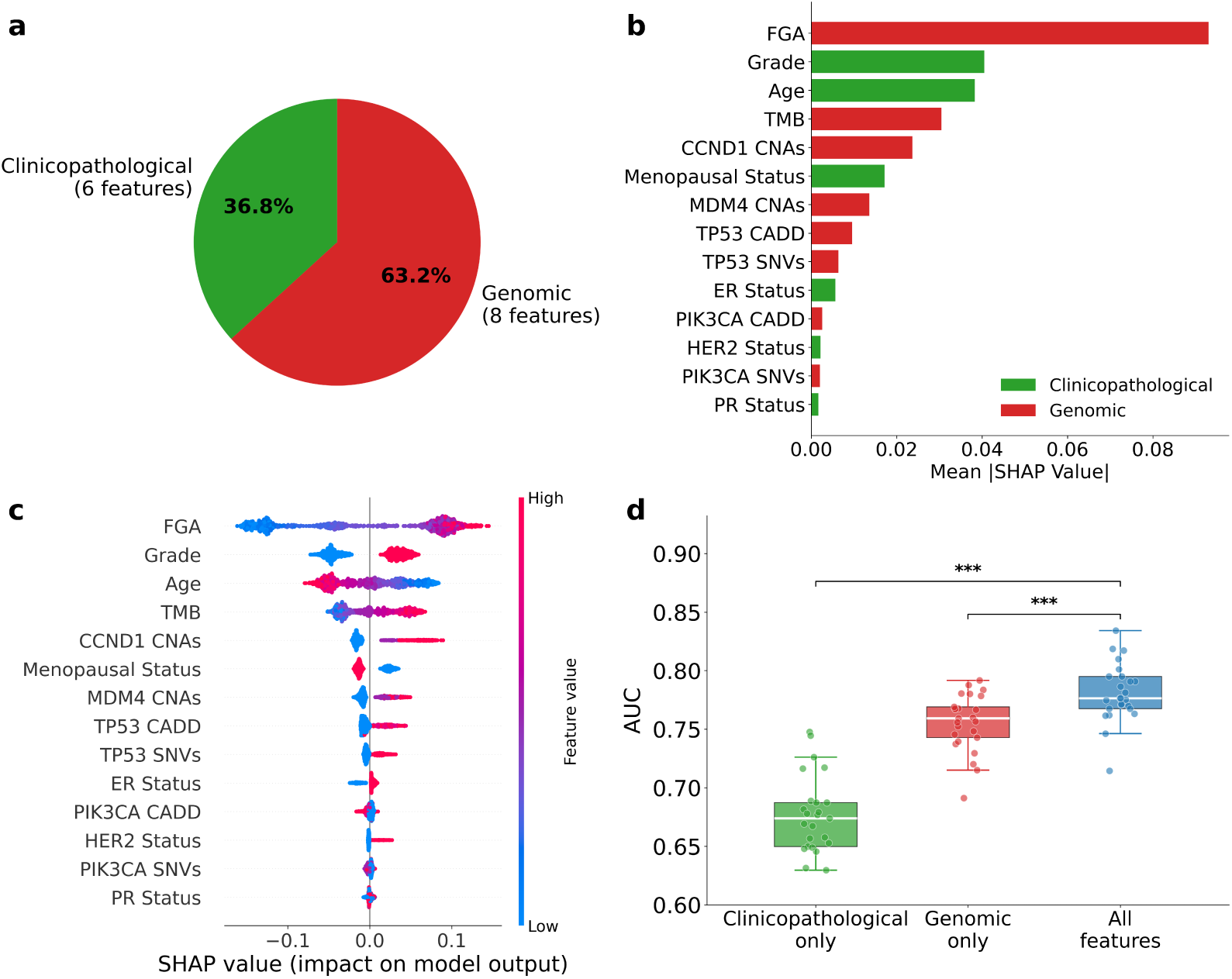
SHAP-based feature importance and ablation analysis. **a** Pie chart showing the proportion of total mean absolute SHAP values attributed to clinicopathological (36.8%) and genomic (63.2%) feature categories. **b** Mean absolute SHAP values for all 14 features ranked by importance. **c** SHAP beeswarm plot showing the direction and magnitude of each feature’s contribution to individual predictions. Each point represents one patient; color indicates feature value (red = high, blue = low). **d** Ablation analysis comparing AUC distributions from repeated 5-fold cross-validation (25 runs) using clinicopathological features only, genomic features only, and the full combined feature set. Statistical significance was assessed using the Wilcoxon signed-rank test; *** p<0.001.

## 4. Discussion

In this study, we developed a multi-cohort machine learning framework for relapse prediction in node-negative stage I-II breast cancer using harmonized clinicopathological and genomic features from MSK-IMPACT and METABRIC cohorts. The model demonstrated moderate-to-good discrimination in bidirectional cross-cohort validation and improved performance in pooled cross-validation. Model-derived risk groups were associated with significant differences in DFS and OS in METABRIC, supporting the prognostic relevance of the learned risk scores. Subtype-stratified analysis further confirmed consistent discrimination across Luminal A, Luminal B, and Triple-Negative subtypes, indicating that the model captures generalizable biological signal rather than subtype-specific patterns. However, because the analysis was retrospective and involved heterogeneous sequencing platforms, differing follow-up structures, and cohort-specific feature distributions, the model should be considered a research-stage prognostic framework requiring prospective validation before clinical decision-support use.

Several machine learning models have been developed for cancer relapse prediction [10, 11, 12]. However, most depend on single cohort validation, where models are trained and evaluated on data from the same institution, making them susceptible to cohort-specific biases. Machine learning algorithms are particularly sensitive to features that provide strong separation within a single cohort [14]. For example, if tumor stage or nodal status strongly discriminates relapse within a particular dataset, the model will adapt to these signals during training, potentially learning cohort-specific patterns rather than generalizable biological relationships. A key finding of this study was the observed asymmetry in cross-cohort generalization performance. Sensitivity analysis revealed that this asymmetry was driven by cohort-specific distributions of age and menopausal status, both of which showed significant differences between relapse and non-relapse groups in MSK-IMPACT but not in METABRIC. When these features were excluded, cross-cohort performance in the train MSK-IMPACT → test METABRIC direction improved across all models, confirming their role as cohort-specific confounders. Although age and menopausal status are established prognostic factors in breast cancer [25], their heterogeneous distributions across cohorts can introduce bias when models are trained on a single institution. Importantly, retaining these features in the combined cohort model led to statistically significant performance gains for RF, GBM, AdaBoost, and XGBoost (p<0.001), demonstrating that when models are exposed to heterogeneous training data from both cohorts, cohort-specific features contribute meaningful signal rather than introducing bias. This finding highlights a critical but underappreciated challenge in biological signals that behave as confounders in a single cohort setting can become informative when the model is trained on sufficiently diverse data. These results underscore the importance of assessing feature distribution differences across cohorts and incorporating sensitivity analyses into cross-cohort validation frameworks.

SHAP analysis revealed that genomic features collectively accounted for 63.2% of the total predictive contribution, with FGA and TMB emerging as the top two genomic features. Notably, both FGA and TMB represent biologically interpretable measures of global genomic instability that have been individually studied as prognostic biomarkers in breast cancer [26]. However, to our knowledge, this study is among the first to harmonize and jointly integrate these features with mutation-level and pathogenicity-weighted genomic variables within a cross-cohort machine learning framework for relapse prediction in node-negative early-stage breast cancer. The selection of TP53 and PIK3CA as the primary genes for SNV and CADD-based features was motivated by their established prognostic significance in breast cancer. TP53 and PIK3CA are the most frequently mutated genes in early relapsed breast cancer [27, 28]. TP53 encodes the guardian of the genome. Wild-type p53 coordinates cell-cycle arrest, apoptosis, and DNA repair in response to genotoxic stress, whereas TP53 mutations impair these protective mechanisms, promoting genomic instability and aggressive tumor behavior [29]. In contrast, PIK3CA mutations predominantly occur in lower-grade HR-positive tumors and paradoxically associate with better short term DFS in this subgroup, likely due to increased dependence on hormone signaling rather than aggressive genomic instability [28]. Consistent with these divergent biological roles, SHAP analysis revealed markedly different contribution patterns for the two genes. TP53 CADD scores and SNV counts showed clear directional positive SHAP values, indicating that high TP53 mutational burden consistently pushed predictions toward relapse. PIK3CA CADD and SNV features, however, showed near-zero and mixed SHAP values, consistent with previous studies suggesting that the prognostic impact of PIK3CA mutations is highly context-dependent, varying by HR status and tumor grade. CCND1 and MDM4 copy number alterations were selected as genomic features based on their established roles in cell cycle dysregulation and p53 pathway suppression respectively. CCND1 amplification leads to cyclin D1 overexpression, which drives aberrant cell cycle progression and has been associated with shorter relapse-free survival in breast cancer [30]. In our SHAP analysis, both CCND1 and MDM4 copy number alterations showed positive SHAP values, with higher copy number consistently pushing predictions toward relapse, consistent with their known oncogenic roles.

Clinicopathological features collectively contributed 36.8% of the total predictive signal, with age and histological grade emerging as the most important features within this category. The positive SHAP values associated with younger age suggest that younger patients are more likely to be classified as high-risk, which is consistent with the well-established observation that younger breast cancer patients tend to present with more aggressive disease. Histological grade showed a clear directional pattern, with higher grade values pushing predictions toward relapse. Menopausal status showed a clear directional pattern, with pre-menopausal status associated with higher relapse risk predictions and post-menopausal status pushing predictions toward non-relapse. This reflects the protective effect of estrogen deprivation in post-menopausal breast cancer. ER, PR, and HER2 status contributed smaller but non-negligible SHAP values, suggesting that while these features provide some prognostic information, their contribution is largely captured by other features in the combined model. Overall, clinicopathological and genomic features provide complementary information. Our ablation analyses demonstrate that significant performance gains when both are combined. Neither feature group alone is sufficient for optimal relapse prediction in node-negative early-stage breast cancer. To support reproducibility and facilitate clinical translation, the final RF model trained on the full combined cohort is publicly available, and all analysis code and results are openly accessible.

Several limitations of this study should be acknowledged. First, while a key strength of this study is the use of bidirectional cross-cohort validation, both MSK-IMPACT and METABRIC are retrospective cohorts. Further evaluation in prospectively collected patient populations is needed to determine the model performance in routine clinical practice. Second, the widely used prognostic tool PREDICT reported moderate discrimination in node-negative patients with an AUC of 0.65 (95% CI: 0.62–0.68) and particularly poor performance in ER-negative tumors (AUC=0.56) [9]. Our model outperforms this benchmark across all experimental settings, achieving an AUC of 0.788 in the combined cohort setting and AUC=0.741 and AUC=0.714 in bidirectional cross-cohort validation. However, a direct head-to-head comparison is not feasible, as PREDICT relies exclusively on clinicopathological variables and treatment information, whereas our model integrates genomic features that are not part of its input parameters. Third, molecular sub-types were defined using IHC-based surrogate criteria based on ER, PR, and HER2 status, as Ki67 data were not uniformly available across both cohorts, which may introduce some misclassifications particularly for Luminal B patients. Fourth, while rank-based inverse normal transformation was applied to harmonize TMB values across MSK-IMPACT and METABRIC platforms, applying this harmonization to external patients sequenced on different panels or in routine clinical settings remains a practical challenge. The transformation parameters are derived from the training cohorts and may not generalize to new institutional data without recalibration. Finally, the study was restricted to node-negative stage I-II patients, and all relapse events in this cohort represent distant metastatic recurrence. The generalizability of the model to lymph node-positive patients or other disease stages therefore remains to be established.

## Supporting information

https://github.com/Cagri-Ozdemir/Early-stage-node-negative-breast-cancer-relapse-prediction/blob/main/Supplementary_Materials.pdf

## Data Availability

All data produced are available online at (https://www.cbioportal.org/).

## 5. Data availability

The preprocessed datasets used in this study are available at https://github.com/Cagri-Ozdemir/Early-stage-node-negative-breast-cancer-relapse-prediction. The original MSK-IMPACT and METABRIC datasets are available at https://www.cbioportal.org/.

## 6. Code availability

The code used for model development, analysis, and experiments is publicly available at https://github.com/Cagri-Ozdemir/Early-stage-node-negative-breast-cancer-relapse-prediction. The software is provided for non-commercial research purposes only.

