## Supplementary material for "Cross-cohort generalizability of machine learning models for early-stage node-negative breast cancer relapse prediction": https://github.com/Cagri-Ozdemir/Early-stage-node-negative-breast-cancer-relapse-prediction/blob/main/Supplementary_Materials.pdf

**Supplementary Table S1.** Variable descriptions and encoding schemes.

| **Variable** | **Description** | **Encoding** |
|---|---|---|
| Age | Age at diagnosis | years / 100 |
| Menopausal status | Pre-menopausal or post-menopausal status at the time of diagnosis | post = 1<br>pre/peri = 0 |
| Histological grade | Tumor differentiation grade (Nottingham system) | well = 1<br>moderate = 2<br>poor = 3 |
| ER status | Estrogen receptor status assessed by immunohistochemistry | positive = 1<br>negative = 0 |
| PR status | Progesterone receptor status assessed by immunohistochemistry | positive = 1<br>negative = 0 |
| HER2 status | Human epidermal growth factor receptor 2 status assessed by immunohistochemistry | positive = 1<br>negative = 0 |
| TMB | Total number of somatic mutations per sample | Continuous:<br>rank-based inverse normal transform |
| FGA | Fraction of genome altered | Continuous:<br>(0–1) |
| TP53 SNVs | Somatic single nucleotide variant counts in TP53 | total SNV count |
| PIK3CA SNVs | Somatic single nucleotide variant counts in PIK3CA | |
| TP53 CADD | Maximum combined annotation dependent depletion pathogenicity score of TP53 variants per sample | maximum CADD score per sample |
| PIK3CA CADD | Maximum combined annotation dependent depletion pathogenicity score of PIK3CA variants per sample | maximum CADD score per sample |
| CCND1 CNAs | Copy number alteration level of CCND1 | Continuous:<br>raw copy number value |
| MDM4 CNAs | Copy number alteration level of MDM4 | Continuous:<br>raw copy number value |

**Supplementary Table S2.** Missing value rates before imputation across the MSK-IMPACT and METABRIC cohorts. Values are reported as the number and percentage of missing samples for each clinical and molecular variable prior to k-nearest neighbor (kNN) imputation.

| Variable | MSK-IMPACT (%) | METABRIC (%) |
|---|---|---|
| Menopausal Status | 7/645 (1.1%) | 69/503 (13.7%) |
| Histological Grade | 104/645 (16.1%) | 18/503 (3.6%) |
| ER Status | 4/645 (0.6%) | 4/503 (0.8%) |
| PR Status | 7/645 (1.1%) | 69/503 (13.7%) |
| HER2 Status | 33/645 (5.1%) | 69/503 (13.7%) |
| FGA | 7/645 (1.1%) | 48/503 (9.5%) |

**Supplementary Table S3.** Optimal hyperparameters for each model under each experimental setting. MSK: MSK-IMPACT cohort; META: METABRIC cohort. Hyperparameters for cross-cohort validations were selected via inner cross-validation on the training cohort only. The final model hyperparameters were determined as the mode of optimal configurations across 25 outer cross-validation runs on the training combined cohort.

| Experiment | Model | Hyperparameters |
|---|---|---|
| Train MSK → Test META | RF | `n_estimators = 100`, `max_depth = 4`, `min_samples_leaf = 3`, `class_weight = 'balanced_subsample'`, `criterion = 'entropy'`. |
| | GBM | `n_estimators = 100`, `max_depth = 3`, `learning_rate = 0.05`, `subsample = 0.7`, `criterion = 'squared_error'`, `loss = 'log_loss'`. |
| | AdaBoost | `n_estimators = 300`, `max_depth = 3`, `learning_rate = 0.05`. |
| | XGBoost | `n_estimators = 200`, `max_depth = 3`, `learning_rate = 0.05`, `subsample = 0.9`, `eval_metric = 'logloss'`. |
| Train META → Test MSK | RF | `n_estimators = 200`, `max_depth = 4`, `min_samples_leaf = 3`, `class_weight = 'balanced_subsample'`, `criterion = 'gini'`. |
| | GBM | `n_estimators = 100`, `max_depth = 4`, `learning_rate = 0.03`, `subsample = 0.5`, `criterion = 'squared_error'`, `loss = 'exponential'`. |
| | AdaBoost | `n_estimators = 200`, `max_depth = 3`, `learning_rate = 0.01`. |
| | XGBoost | `n_estimators = 300`, `max_depth = 4`, `learning_rate = 0.01`, `subsample = 0.7`, `eval_metric = 'logloss'`. |
| Combined Cohort (The Final Model) | RF | `n_estimators = 300`, `max_depth = 4`, `min_samples_leaf = 7`, `class_weight = 'balanced'`, `criterion = 'entropy'`. |
| | GBM | `n_estimators = 300`, `max_depth = 3`, `learning_rate = 0.01`, `subsample = 0.5`, `criterion = 'squared_error'`, `loss = 'exponential'`. |
| | AdaBoost | `n_estimators = 200`, `max_depth = 3`, `learning_rate = 0.05`. |
| | XGBoost | `n_estimators = 300`, `max_depth = 3`, `learning_rate = 0.01`, `subsample = 0.5`, `eval_metric = 'logloss'`. |

**Supplementary Table S4.** Predictive performance of machine learning models for breast cancer relapse prediction across experimental settings. Metrics include area under the receiver operating characteristic curve (AUC), area under the precision–recall curve (AUCPR), recall, precision, F1-score, and specificity. For the combined cohort setting, results are reported as mean ± standard deviation across 5-fold cross-validation repeated five times (25 runs). MSK: MSK-IMPACT cohort; META: METABRIC cohort.

| Model | Experiments | AUC | AUCPR | Recall | Precision | F1 | Specificity |
|---|---|---|---|---|---|---|---|
| | Train MSK → Test META | 0.714 | 0.663 | 0.720 | 0.616 | 0.664 | 0.616 |
| RF | Train META → Test MSK | 0.741 | 0.689 | 0.722 | 0.669 | 0.694 | 0.657 |
| | Combined cohort (5-fold) | 0.781±0.024 | 0.758±0.029 | 0.721±0.053 | 0.693±0.030 | 0.706±0.033 | 0.707±0.042 |
| | Train MSK → Test META | 0.669 | 0.603 | 0.634 | 0.579 | 0.605 | 0.605 |
| GBM | Train META → Test MSK | 0.700 | 0.646 | 0.604 | 0.645 | 0.624 | 0.681 |
| | Combined cohort (5-fold) | 0.785±0.025 | 0.775±0.029 | 0.693±0.055 | 0.711±0.033 | 0.701±0.035 | 0.741±0.043 |
| | Train MSK → Test META | 0.674 | 0.627 | 0.625 | 0.585 | 0.604 | 0.620 |
| AdaBoost | Train META → Test MSK | 0.717 | 0.675 | 0.551 | 0.642 | 0.593 | 0.705 |
| | Combined cohort (5-fold) | 0.784±0.026 | 0.772±0.031 | 0.692±0.062 | 0.703±0.034 | 0.696±0.037 | 0.732±0.047 |
| | Train MSK → Test META | 0.668 | 0.631 | 0.578 | 0.604 | 0.590 | 0.675 |
| XGBoost | Train META → Test MSK | 0.688 | 0.631 | 0.522 | 0.632 | 0.572 | 0.708 |
| | Combined cohort | 0.788±0.026 | 0.778±0.028 | 0.691±0.059 | 0.711±0.039 | 0.700±0.039 | 0.742±0.048 |

**Supplementary Fig. S1.** Subtype-stratified Kaplan-Meier survival curves. Disease-free survival (DFS; **a**–**c**) and overall survival (OS; **d**–**f**) for Luminal A, Luminal B, and Triple-Negative subtypes, respectively. Risk groups were defined using a probability threshold of 0.5 based on out-of-fold predictions from the combined cohort cross-validation. Shaded regions represent 95% confidence intervals.

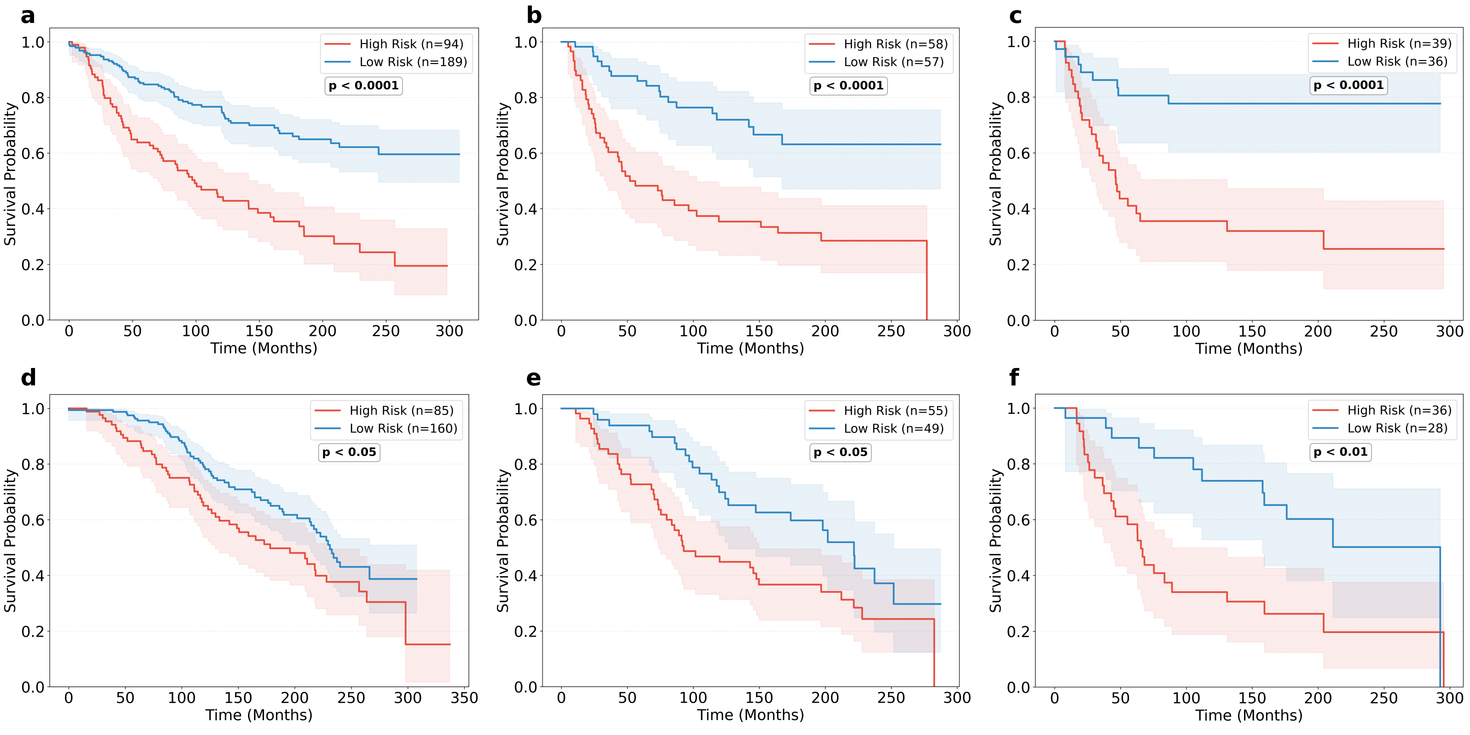

**Supplementary Fig. S2.** Decision plots illustrating model-derived relapse risk predictions for two representative patients: **a** high-risk patient prediction. **b** low-risk patient prediction. Each horizontal line represents the cumulative contribution of a feature to the final risk probability, starting from the baseline of 0.5. Red segments indicate features pushing predictions toward relapse (positive SHAP values) and blue segments indicate features pushing predictions toward non-relapse (negative SHAP values). Feature values for each patient are shown on the left. The final prediction f(x) is indicated by the vertical line.

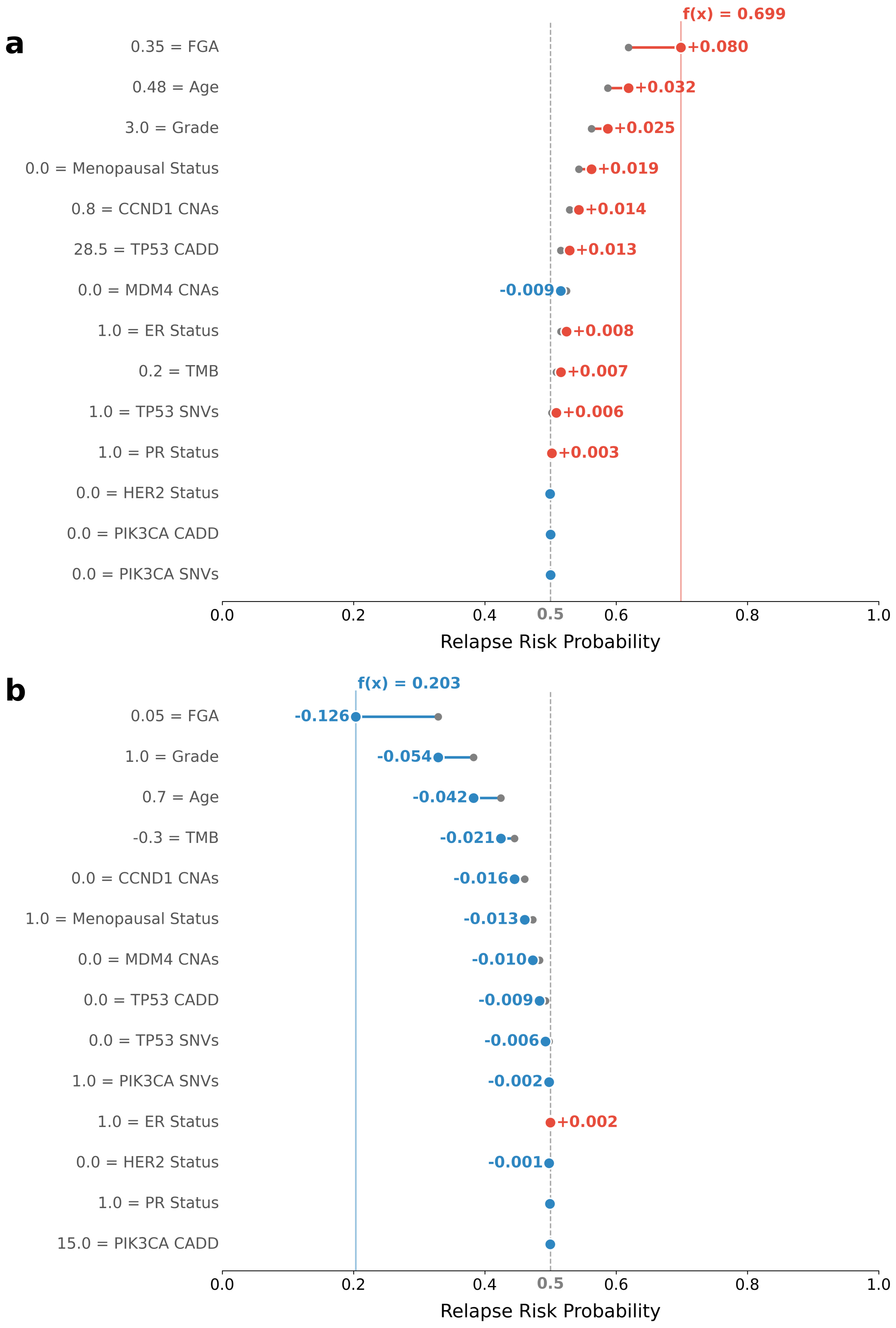
